# Cross-modal Functional Plasticity after Cochlear-implantation

**DOI:** 10.1101/2024.08.22.24312200

**Authors:** Jamal Esmaelpoor, Tommy Peng, Beth Jelfs, Darren Mao, Maureen J. Shader, Colette M. McKay

**Author notes:** Corresponding author *Email address:* (Jamal Esmaelpoor).

## Abstract

**Objective:** Despite evidence that cross-modal effects after hearing loss and cochlear implantation are primarily conveyed through synaptic gain and efficacy rather than reorganized fiber tracts, few studies have assessed cross-modal functional connectivity (CMFC) to evaluate plasticity. This study, inspired by the psychophysiological interactions (PPI) method, addresses its limitations and provides a robust approach to evaluating task-induced CMFC.

**Design:** Twenty-two post-lingually deafened, newly implanted adult cochlear implant (CI) recipients with severe hearing loss in the contralateral ear and 17 normal-hearing (NH) subjects participated. The experiment included audio-only and visual-only speech tasks, with resting-state FC as a baseline. Functional near-infrared spectroscopy (fNIRS) measured brain imaging data one month and one year post-implantation. CI users’ speech understanding performance was evaluated one year after implantation.

**Results:** A negative correlation was found between average contralateral task-induced CMFC and speech outcomes, particularly in links from the angular gyrus (AG), both one month and one year post-activation. Plastic changes showed higher task-induced CMFC in AG compared to the superior temporal gyrus (STG), aligning with neural efficiency principles. Task-induced CMFC remained elevated in CI users compared to NH cohorts even after one year.

**Conclusion:** Task-induced CMFC can serve as a significant marker of cross-modal plasticity and speech performance in CI recipients, indicating increased reliance on cross-modal processing in one year after implantation.

## 1. Introduction

Loss of a sensory modality, such as hearing in cases of post-lingual hearing loss, causes a phenomenon known as cross-modal plasticity in the brain. Cross-modal plasticity is characterized by the need to compensate for the deprived sensory modality resulting in heightened brain activity in regions associated with the remaining senses. Consequently, the primary cortical area previously dedicated to the lost modality undergoes a takeover by other senses [1, 2]. Notably, individuals with severe hearing loss often adapt by relying more on intact senses such as vision. In these cases, the auditory cortex becomes increasingly responsive to visual stimuli, enhancing capabilities in visual object localization and motion detection [3, 4, 5].

Cochlear implants offer a partial restoration of hearing ability, for individuals with severe hearing loss, through electrical stimulation of ganglion cells in the auditory nerves. The introduction of auditory stimulation post-cochlear implantation (CI) triggers cross-modal plasticity in visual and auditory cortices, enabling the brain to adapt to the novel stimuli. However, post-CI plasticity is complex and multifaceted. First, sensorineural hearing loss precipitates various changes, including a significant reduction in spiral ganglion neurons, demyelination of residual neurons, shrinkage of perikaryon in the auditory pathway, and a decrease in spontaneous activity in the auditory pathway [6]. Second, the implant delivers signals with coarse spectral content compared to natural acoustic hearing [7]. Consequently, post-lingual implant recipients must forge new associations with the sounds received after implantation, often necessitating additional neural resources, such as increased cognitive load and continued reliance on supplementary cues like lipreading [8, 9].

Early animal model studies have demonstrated that when early deafness is combined with the removal of the auditory midbrain, it results in the reorganization of anatomical inputs to the thalamus and a large-scale remapping of the auditory cortex by visual inputs [10, 11]. Influenced by this perspective, many studies suggested that the entire auditory cortex becomes a battleground for sensory systems, with each area potentially being recruited for a new sensory function in the absence of auditory input [12, 13]. However, recent findings challenge this notion, refuting the idea that massive cross-modal reorganization occurs in the auditory system after hearing loss without anatomical rerouting of visual information. These findings suggest that the cause of cross-modal reorganization is primarily synaptic and functional, confined to restricted auditory areas. The brain can repurpose the same neuronal circuitry for qualitatively different functions [5]. Cross-modal effects are conveyed predominantly through synapses, their number, and synaptic efficacy rather than reorganized fiber tracts. Therefore, assessing functional connectivity (FC), which involves evaluating statistical dependencies between activities in different brain areas [14], appears to be a reasonable approach for understanding changes related to cross-modal reorganization.

Even though cross-modal regional activation has been widely assessed by many studies [3, 4, 15, 16, 17, 8], there are only a limited number of studies that have explored cross-modal functional connectivity (CMFC) for post-cochlear implantation users. These studies on CMFC exhibit disparities in their conceptualizations of FC and the specific aspects they evaluate. For example, Chen et al. investigated task-induced CMFC between the visual and auditory cortices during both visual and auditory tasks among CI users and individuals with normal hearing (NH) [18]. Employing functional near-infrared spectroscopy (fNIRS) for brain imaging, they selectively excluded channels demonstrating the highest activation during visual and auditory tasks in their respective areas. This selection criterion stemmed from the presumption that cross-modal changes predominantly occur in secondary areas. Additionally, they normalized CMFC, measured via Pearson correlation, by subtracting average intramodal connectivity between channels in the left and right hemispheres, assuming negligible task effects on them and regional disparities in baseline FC. The outcomes revealed heightened task-induced CMFC in CI users compared to the normal hearing group. Moreover, the discrepancy in CMFC for visual and auditory stimuli correlated with speech recognition outcomes in CI users.

In contrast, Fullerton et al. focused on channels exhibiting the highest task activation for CMFC analyses, also using fNIRS for brain imaging [9]. They quantified FC between channels using coherence, focusing on the raw values of FC between channels. This approach potentially assumes minimal residual physiological noise after band-pass filtering, or negligible individual/regional differences in physiological noise, which given the characteristics of fNIRS signals would be strong assumptions [19]. Their findings highlighted a positive correlation between connectivity in the left auditory and visual cortices and CI users’ abilities to comprehend speech in background noise. However, they overlooked the transient effects of tasks when evaluating FC during the task. These divergent methodologies underscore the complexities and variations inherent in investigating cross-modal FC within the realm of auditory rehabilitation.

An alternative method for investigating task-specific changes is Psychophysiological Interactions (PPIs) analysis, introduced by Friston et al. [20]. PPI assesses changes in the relationship between activity in different brain areas using a regression model with three regressors for each seed channel. The main regressor of interest is the psychophysiological interaction, however, to obtain this we must first consider the interactions which are either purely pschological or purely physiological. To rule out the transient effect of the task itself (psychological interaction), a regressor is obtained by convolving a boxcar function representing the duration of the task with the hemodynamic response function (HRF) to give the HRF-convolved task. To account for the anatomical connections with the seed region (physiological interaction), the time course from the seed region of interest is used as a regressor. This accounts for the correlations present all the time, both during and outside of the task duration. The final regressor, is the element-wise product of the HRF-convolved task (black line) and seed channel. This covariate represents the psychophysiological interaction and accounts for the changes in FC due to the task [21]. The generalized form of PPI for scenarios including two tasks examines the difference in effects of different tasks on baseline FC and does not provide their effects separately [22]. Therefore, this study aims to address the gap in the literature by introducing a reliable approach for assessing task-induced CMFC.

### 1.1. Aims and Contributions

This study investigates task-induced CMFC in post-lingually deaf CI users, focusing on the interaction between visual and auditory brain regions during visual and audio tasks and its correlation with speech comprehension one year after cochlear implantation. fNIRS, a non-invasive imaging technique that measures cortical brain activity by detecting changes in blood oxygenation and blood volume without interfering with the CI device [23], was used for recordings at two time points: one month and one year post-implantation. To achieve these aims, we devised an approach inspired by PPI. However, we acknowledge that utilizing PPI itself imposes certain limitations. To address these constraints, we opted for resting-state recordings instead of short intervals between task trials (control trials) to ascertain baseline FC and to deal with the restriction of generalized PPI for scenarios including two tasks [22].

Our primary hypothesis is that task-induced CMFC correlates with the behavioral speech outcomes of CI users. Additionally, we undertake further analyses to evaluate differences in cross-modal plasticity in the primary and secondary auditory areas covered by our montage without making any presumptions about this debated issue in previous studies. Lastly, we compare CMFC in CI recipients with that of a NH group to identify potential disparities.

## 2. Materials and Methods

### 2.1. Participants

We enrolled twenty-six post-lingually deafened newly implanted adult CI recipients with severe hearing loss in the contralateral ear (pure-tone average (PTA) of 0.5, 1, 2, and 4 kHz *>* 45 dBHL) for this study. As the study evaluates task-induced changes in FC with resting-state serving as the baseline, four subjects were excluded due to missing resting-state recordings. Consequently, data from twenty-two CI recipients were utilized (mean age = 58*±*14 years, 13 male, 9 female; demographic information in Table 1). Additionally, 17 NH subjects with PTA *<* 45 dBHL in both ears participated in the experiment (mean age = 58*±*14 years, 8 male, 9 female). This study was approved by the Human Research Ethics Committee of the Royal Victorian Eye and Ear Hospital, Melbourne, Australia (ethics approval 19.1418H). All participants provided informed written consent.

**Table 1:** Demographic Information of Participants.

| Participant | Gender | Age<br>[Range] | Implant Ear | Deafness<br>Duration<br>[Years] | Non-Implant<br>Ear PTA*<br>[dBHL] |
| --- | --- | --- | --- | --- | --- |
| Sub-01 | M | 61-65 | R | N.R.<sup>†</sup> | 64 |
| Sub-02 | F | 46-50 | R | 20 | 56 |
| Sub-03 | F | 55-60 | R | 48 | 76 |
| Sub-04 | M | 71-75 | R | 60 | 92 |
| Sub-05 | F | 71-75 | R | 30 | 111 |
| Sub-06 | M | 61-65 | R | 30 | 66 |
| Sub-07 | M | 26-30 | L | 10 | 106 |
| Sub-08 | M | 61-65 | L | 27 | 72 |
| Sub-09 | M | 61-65 | L | 15 | 74 |
| Sub-10 | M | 41-45 | R | 30 | 69 |
| Sub-11 | F | 41-45 | R | 0.17 | 87 |
| Sub-12 | M | 31-35 | L | 7 | 101 |
| Sub-13 | M | 56-60 | R | 45 | 65 |
| Sub-14 | M | 66-70 | R | 43 | 120 |
| Sub-15 | F | 66-70 | L | N.R. | 97 |
| Sub-16 | F | 66-70 | R | 28 | 76 |
| Sub-17 | F | 61-65 | R | 25 | 81 |
| Sub-18 | M | 71-75 | R | 30 | 104 |
| Sub-19 | M | 31-35 | R | 25 | 71 |
| Sub-20 | F | 61-65 | L | 30 | 96 |
| Sub-21 | M | 71-75 | R | 20 | 80 |
| Sub-22 | F | 71-75 | R | 8 | 67 |

\* PTA (dBHL) = pure-tone average of 0.5 1 and 2 4 kHz hearing thresholds.
<sup>†</sup> N.R. = no record

Given the older age group of our participants, their cognitive skills were assessed to identify any potential cognitive decline. Cognitive skills were evaluated using trial-making tests A and B [24]. Test A required participants to draw lines between numbers in ascending order, while Test B involved drawing lines alternately between numbers and letters in ascending order. Both tests were performed on a sheet of paper with random placements. Participants were instructed to complete the tasks quickly and accurately without lifting the pencil. All participants demonstrated normal performance, meeting the specified criteria [25].

CI outcomes were assessed one year after device activation. Participants underwent two audio-only speech tests to measure speech understanding performance. The first test involved 50 consonant-nucleus-consonant (CNC) words presented in quiet [26]. The second test comprised 15 Bamford-Kowal-Bench (BKB) sentences presented in four-talker babble noise (at a signal-to-noise ratio (SNR) = 10 dB) [27]. Participants were seated comfortably and experienced free-field presentation of speech stimuli at 65 dBA through their implants. To isolate the participant performance with the implant, the contralateral ear was occluded and masked when appropriate. Test scores for the first test were based on the percentage of correct phonemes recognized, while for the second test it was the percentage of correct words in repeated sentences. The speech score results were transformed into rationalized arcsine units for statistical analysis [28].

### 2.2. Experiments

We conducted a procedure comprising a task-based experiment with audio and visual tasks followed by a resting-state recording as outlined in [29]. Participants were seated in a comfortable chair within a sound-attenuated booth facing a monitor screen. A loudspeaker was positioned 1 meter from the participant for the free-field presentation of auditory stimuli at 55 dBA.

The experiment was comprised of a task-based component followed by a resting-state component. During the task-based component (Fig. 1(a)), participants attended to 36 contextually connected segments of a children’s story (”Mrs. Tittlemouse” by Beatrix Potter) presented by a female narrator. The story segments were either audio-only or visual-only and had an average length of 12.5 seconds. In audio-only trials, participants heard the story through the speaker with a fixation gray cross on the screen. During visual-only speech trials, the participants watched a video of a female narrator tell the story without sound. Furthermore, during 10 control segments, the participants attended to a gray fixation cross on the screen with no sound. All segments were followed by 15-30 second intervals during which the participants attended to a gray fixation cross on the screen with no sound. Segments were pseudo-randomized in condition while keeping the segments contextually continuous. After the task-based component, we conducted a 5-minute closed-eyes resting-state recording. Participants were instructed to close their eyes and relax during the recording.

**Figure 1:**
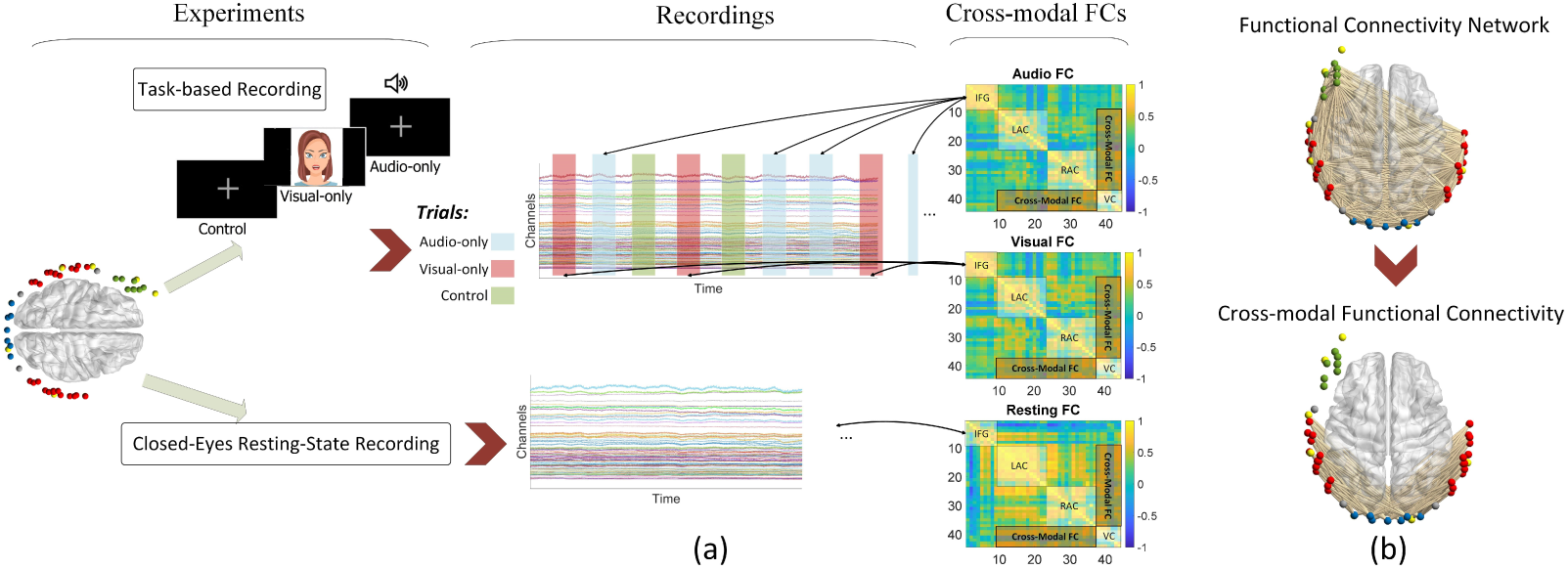
(a) Experimental design overview: The experiment comprises two steps. The first step is task-based, involving participants listening to a short story with audio-only, visual speech-only, and control trials. The second step involves a 5-minute closed-eye resting state. Based on the recordings from these two steps, adjacency matrices (FC matrices) are calculated for both resting state and task trials. The resting-state FC serves as the baseline to measure task-related components in FC. (b) Cross-modal functional connectivity links: Illustration of the cross-modal functional connectivity links investigated in this study, depicting connections between auditory and visual cortices.

### 2.3. fNIRS Recording

Two fNIRS recordings were conducted in this study: one within the first month after implant switch-on and the second at one year after implant switch-on. We employed the same fNIRS acquisition setup for both recordings. A continuous-wave NIRScout device (NIRScout, NIRX medical technologies, LLC) was used for data acquisition. The montage comprised 16 sources and 16 photodiode detectors. The sources included two near-infrared light emitters illuminating at wavelengths of 760 and 850 nm. This configuration established 44 long-channels with an average separation of 3 cm between sources and detectors. The channel placements, as depicted in Fig. 2, covered the inferior frontal gyrus (green channels), right and left auditory cortices (red channels), and occipital lobe (blue channels) cortical areas of the brain. Additionally, three channels were positioned between these areas, indicated in grey in the figure. The montage also featured 8 short-channels (yellow channels) with an 8 mm separation between their sources and detectors.

**Figure 2:**
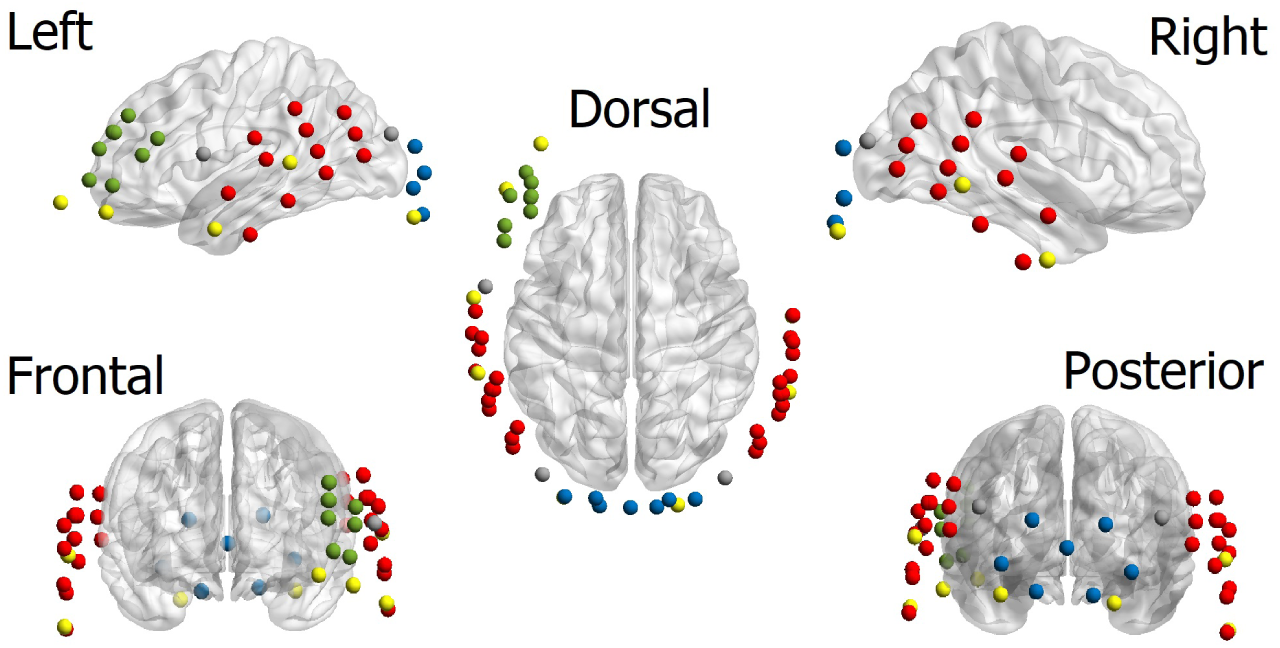
Data Acquisition Montage: The montage comprises 44 long channels with approximately 3 cm distance between sources and detectors, and 8 short channels with an 8 mm separation between sources and detectors. The montage covers the left inferior frontal gyrus (green channels), left and right auditory cortices (red channels), and the occipital lobe (blue channels). Short channels are highlighted in yellow.

### 2.4. Data Preprocessing

The MATLAB software, along with the NIRS toolbox [30], was employed for data preprocessing. Initially, fNIRS recordings were converted to optical density. Channel quality was assessed based on the scalp coupling index (SCI) [31], with a threshold of 0.5 defining bad channels. Channels with SCI values below 0.5 were excluded from subsequent analysis. The temporal derivative distribution repair (TDDR) method was then applied to the remaining channels to enhance signal quality by mitigating motion artifacts [32]. Subsequently, the optical signals were converted to oxy- and de-oxyhemoglobin (HbO and HbR) using the modified Beer-Lambert Law [33]. Given that long channels capture both cerebral and systemic components and artifacts such as heartbeats, respiration, and Mayer waves, two successive steps of short-channel correction and band-pass filtering were implemented to mitigate the impact of these artifacts. Short-channel correction involved regressing out short channels (containing systemic artifacts and no cerebral component) from long channels [19]. An FIR band-pass Butterworth filter with an order of 8 and a band-pass range of 0.033 to 0.40 Hz was applied to remove low-frequency artifacts like baseline drifts and high-frequency artifacts like heartbeats. Finally, a general linear model analysis (GLM) was performed to mitigate transient effects of tasks using canonical HRF-convolved task regressor, akin to the approach employed by PPI method [21, 34].

### 2.5. Cross-modal Functional Connectivity

As detailed, the study aims to evaluate task-induced CMFC with resting state as the baseline and its correlation with the behavioral speech performance of cochlear implant (CI) users. FC adjacency matrices were computed for tasks involving audio-only and visual-only trials, as well as during resting state. Pearson correlation was utilized to compute FC, with the application of the Fisher transform to enhance the normality of weight distributions [35]. FC was calculated during the 30-second intervals of audio-only and visual-only tasks (Fig.1 1(a)) and then normalized by subtracting resting-state FC from them. To mitigate spurious components in FC, the lower cut-off frequency for the band-pass filter in the preprocessing step was set to 1/30 *≈* 0.033 Hz [36, 37]. The study specifically focuses on cross-modal FC links, defined as links connecting nodes in the auditory cortex to nodes in the visual cortex (Fig.1(b)).

## 3. Results

One of the key objectives of this study was to assess cross-modal plasticity in the primary and secondary auditory cortices concerning task-induced CMFC. To achieve this, we adopted the auditory areas classification in our montage proposed by [29]. Shader et al. employed the fNIRS Optode Location Decider (fOLD) tool to partition the auditory region into two subregions: superior temporal gyrus (STG) and angular gyrus (AG) ^1^, representing distinct anatomical structures of the primary and secondary auditory areas, respectively.

### 3.1. Task-induced CMFC and CI Outcomes: Broad Auditory Areas

Motivated by the fact that our participants only hear through the implant side, we examined the correlation between average contra- and ipsilateral CMFC links, relative to the implantation side, and behavioral speech score outcomes across all channels within the auditory cortex. Significant negative correlations were found between average contralateral task-related CMFC during the first fNIRS recording (one month after device activation) and CNC phoneme scores in quiet during both visual-only and audio tasks (Fig.3). All p-values were corrected for multiple comparisons using the Bonferroni method, multiplying the p-values by 4. In the main text, we present correlation results exclusively for contralateral links, as all results for ipsilateral links were insignificant.

**Figure 3:**
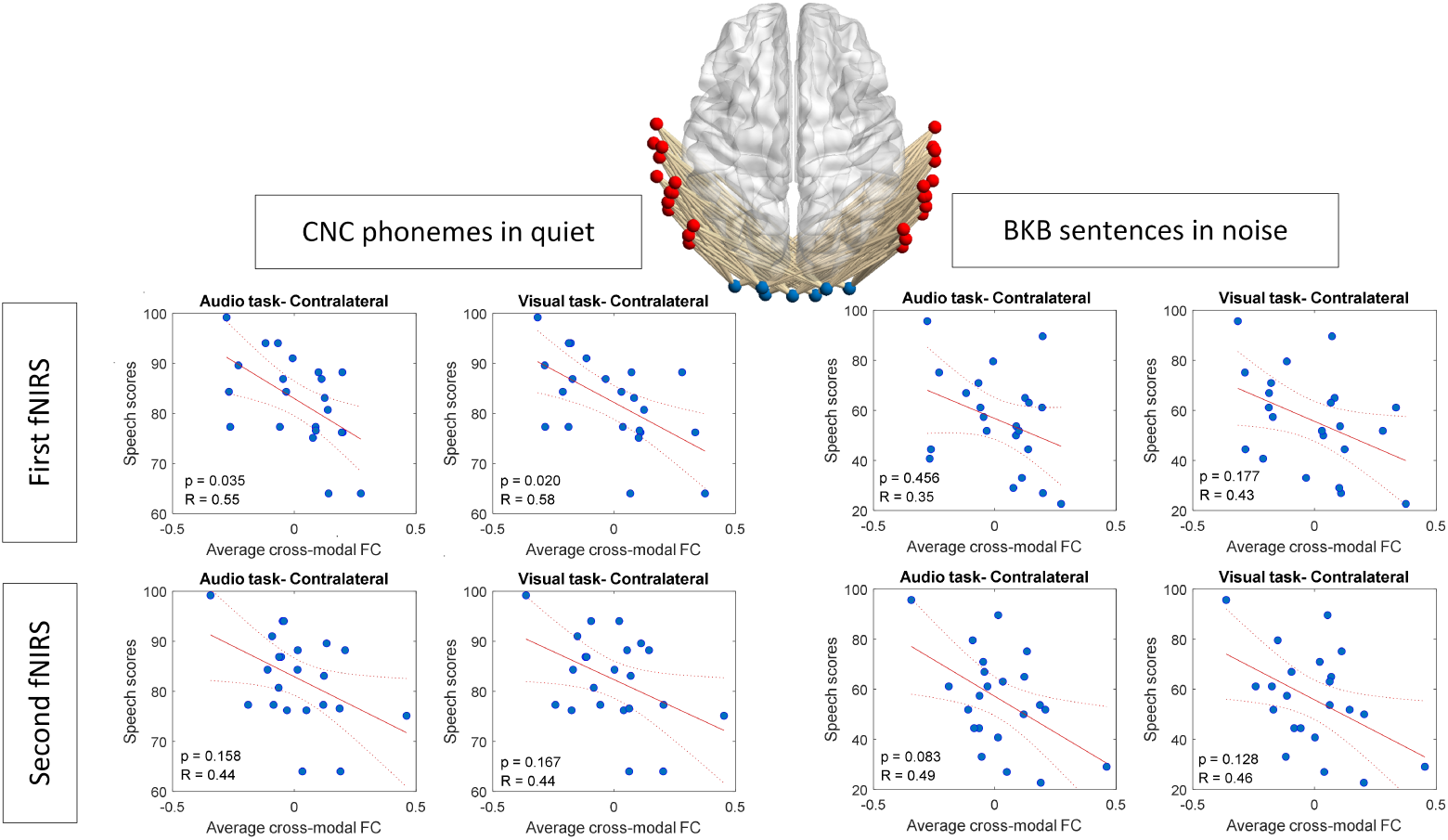
Correlation between average contra-lateral cross-modal functional connectivity (FC) and speech outcomes, employing all channels in the auditory cortex (AC) representing the broad auditory area.

### 3.2. Task-induced CMFC and CI Outcomes: Restricted Auditory Areas

Next, focusing on the restricted auditory regions of the angular gyrus (AG) and superior temporal gyrus (STG) separately, we examined the correlation between average task-related CMFC and the speech performance outcomes of CI recipients. As illustrated in Fig.4 and Fig.5, the correlations between speech scores and task-induced CMFC for contralateral links were predominantly significant and negative for links originating from AG to VC. However, the significance of these correlations diminished when considering links from STG to VC. All p-values were corrected for multiple comparisons using the Bonferroni method, multiplying the p-values by 8.

**Figure 4:**
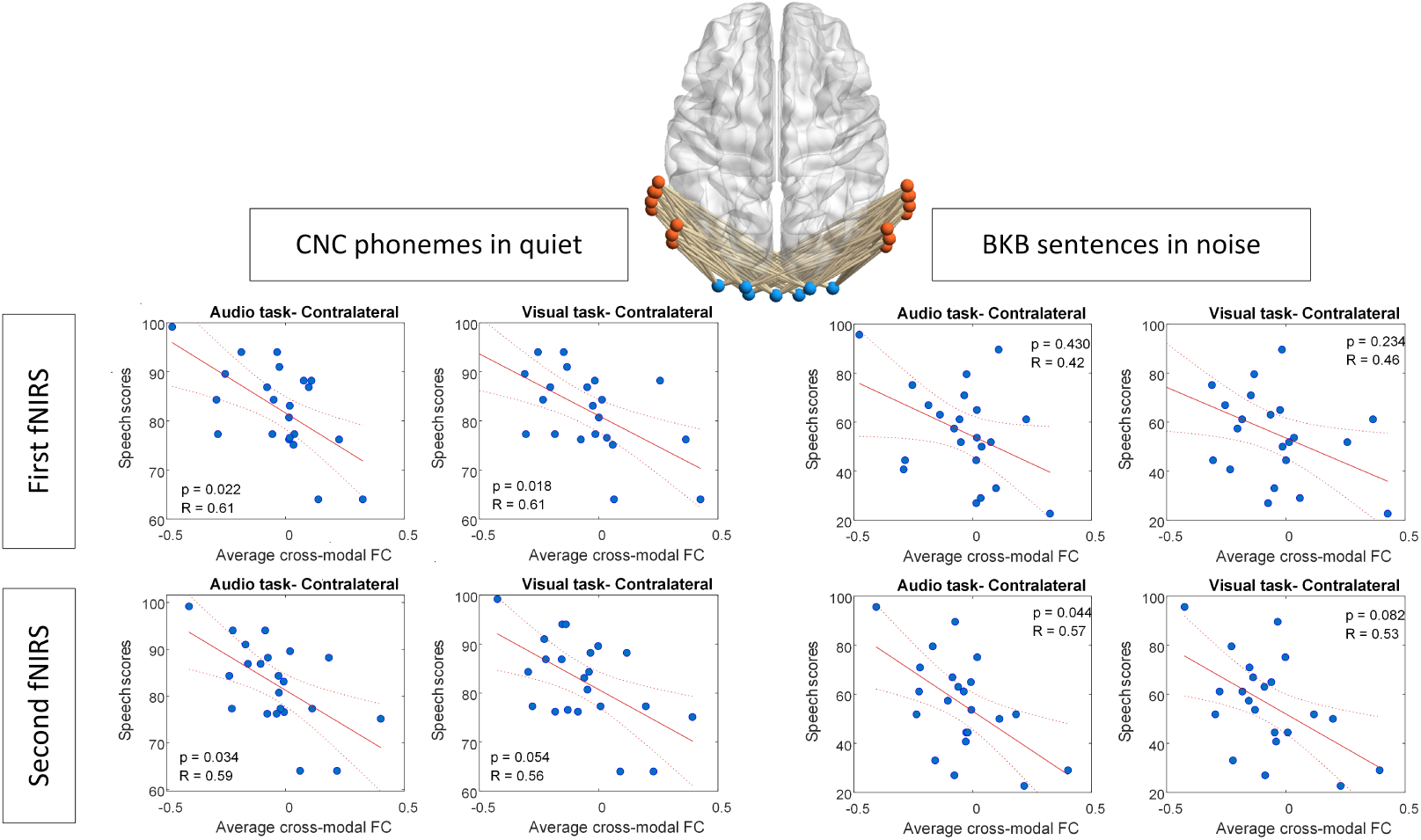
Correlation between average contra-lateral task-induced CMFC and speech outcomes, utilizing FC links from channels in AG (indicated in pink color) to channels in the VC (in blue color).

**Figure 5:**
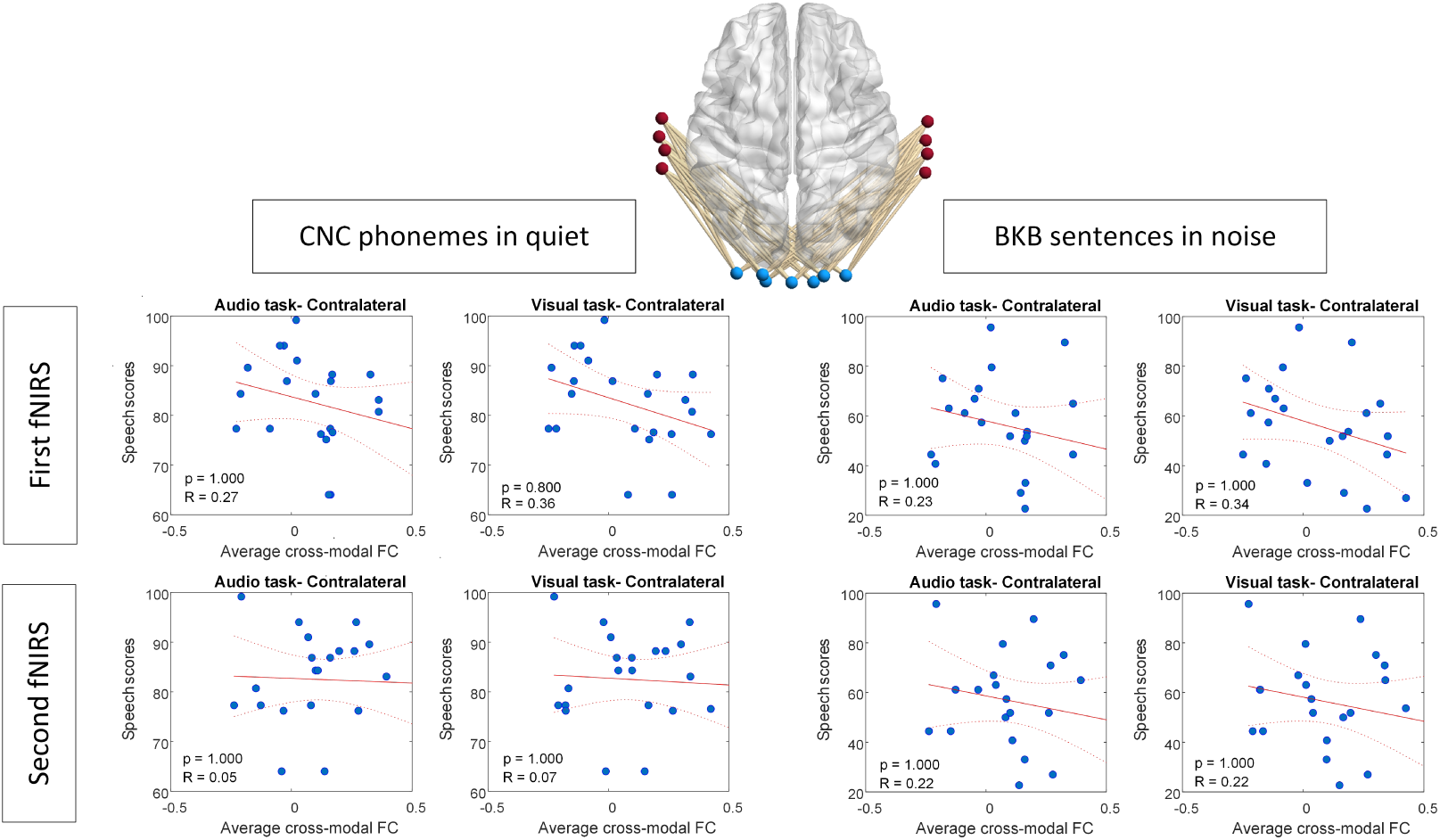
Correlation between average contra-lateral task-induced CMFC and speech outcomes, utilizing FC links from channels in STG (indicated in maroon color) to channels in the VC (in blue color).

### 3.3. Cross-modal Link Importance Based on CI Outcomes

To better understand the distinction between cross-modal links in AG and STG concerning their correlation with speech performance outcomes one year after CI activation, we employed univariate feature ranking for regression [38]. This analysis enabled us to determine the relative significance of individual links in predicting performance outcomes. Using F-tests, we assessed the importance of each link in estimating speech performance outcomes, with the negative logarithm of the p-values serving as the scores (implemented using the fsrftest function in MATLAB). Consistent with our findings based on average link weights in different restricted areas, and by comparing significance scores for groups of links from AG and STG to the VC through unpaired two-sample t-tests, the results underscore the higher significance of links from AG in both fNIRS recordings concerning correlation with speech understanding performance outcomes (Fig.6).

**Figure 6:**
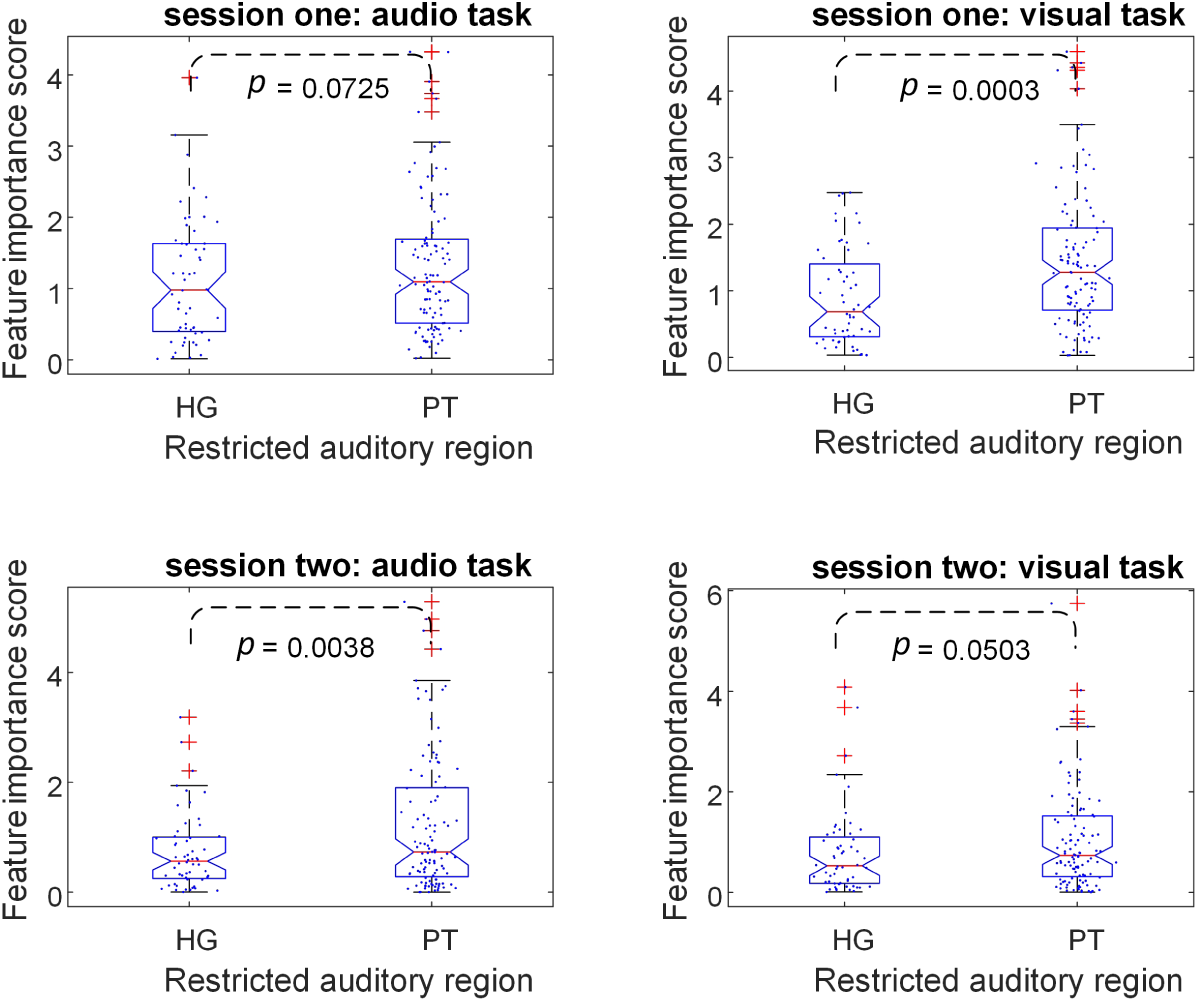
Significance of individual links in predicting performance outcomes in STG and AG. The relative importance was determined using univariate feature ranking for regression via F-tests. Links from AG to VC show a stronger correlation with the behavioral speech performance of CI users compared to those from STG. Dashed lines represent two-group comparisons.

### 3.4. Comparing Task-induced CMFC of STG and AG after CI

In addition to assessing the correlation between links from AG and STG and behavioral speech performance outcomes, we compared these weights for the two fNIRS recording sessions conducted at one month and one year post-switch-on. Using paired t-tests, Fig.7 compares the average cross-modal link weights from AG and STG to VC for CI recipients in both recording sessions and various tasks. The results indicate that while the difference between average CMFC links was not significant one month post-implant switch-on, it became significantly different after one year, suggesting a higher task-induced CMFC for the AG area.

**Figure 7:**
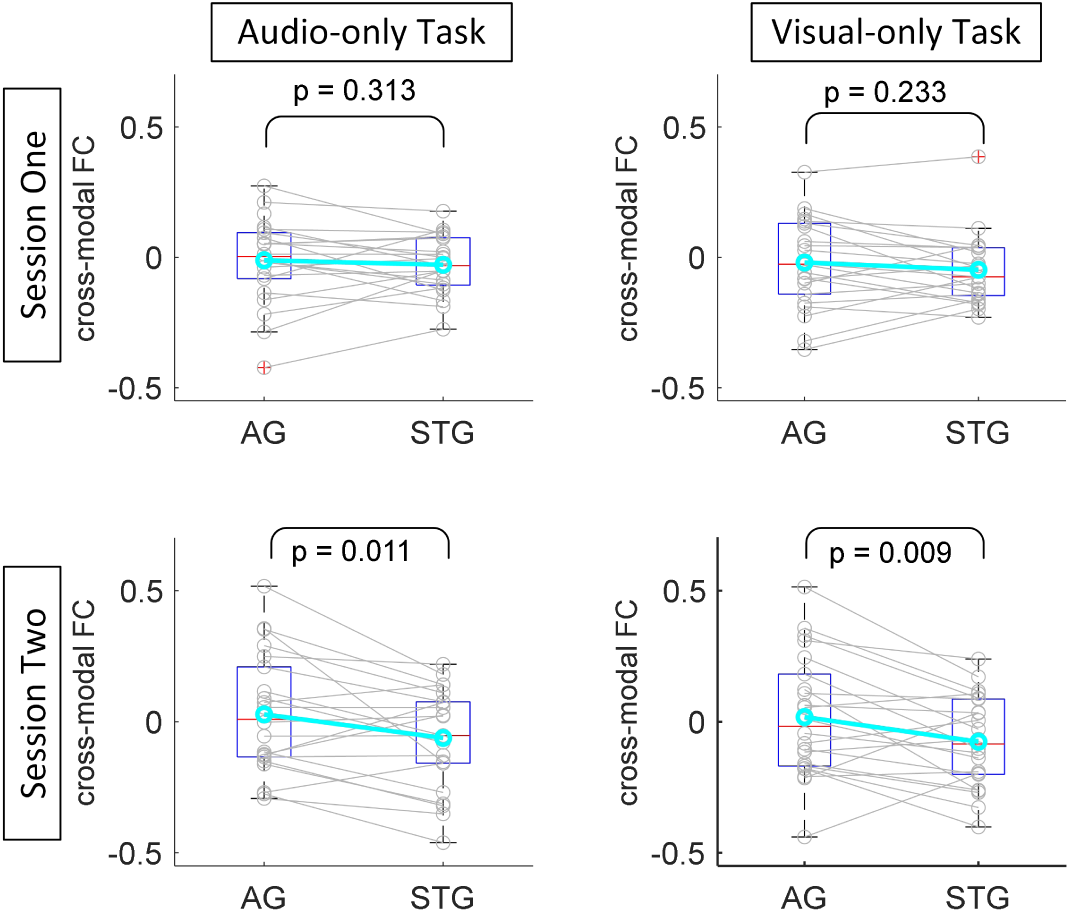
Comparison of average task-induced CMFC between AG and STG links to VC in CI recipients. Initially insignificant shortly after implantation (one month post device switch-on), but after one year, task-induced CMFC becomes significantly higher for AG links. Solid lines represent paired t-tests.

### 3.5. Task-induced CMFC and Channel Pair Distances

Given the notable differences observed in task-induced CMFC of links connecting AG and STG to VC in the results discussed so far, we sought to investigate the relationship between link length and task-induced CMFC. The rationale behind this exploration was the apparent difference between AG and STG in terms of their distance to VC (STG channels being farther away from VC compared to channels in AG). Therefore, we aimed to examine the impact of link length on the task-induced CMFC. In this section, we focused on those links that connect channels in each hemisphere but not across hemispheres. We conducted the same analysis for both NH subjects and CI recipients. As illustrated in Fig.8, the average changes in task-induced CMFC across subjects negatively correlate with channel pair distance in the NH population. For CI users, our results indicate a weaker correlation shortly after implantation (one month after CI switch-on), while after one year, a similar strong negative correlation to the NH group emerges between task-induced CMFC and distance. This implies that topologically closer areas cooperate more (show more correlated activities) to accomplish the audio- and visual-only tasks.

**Figure 8:**
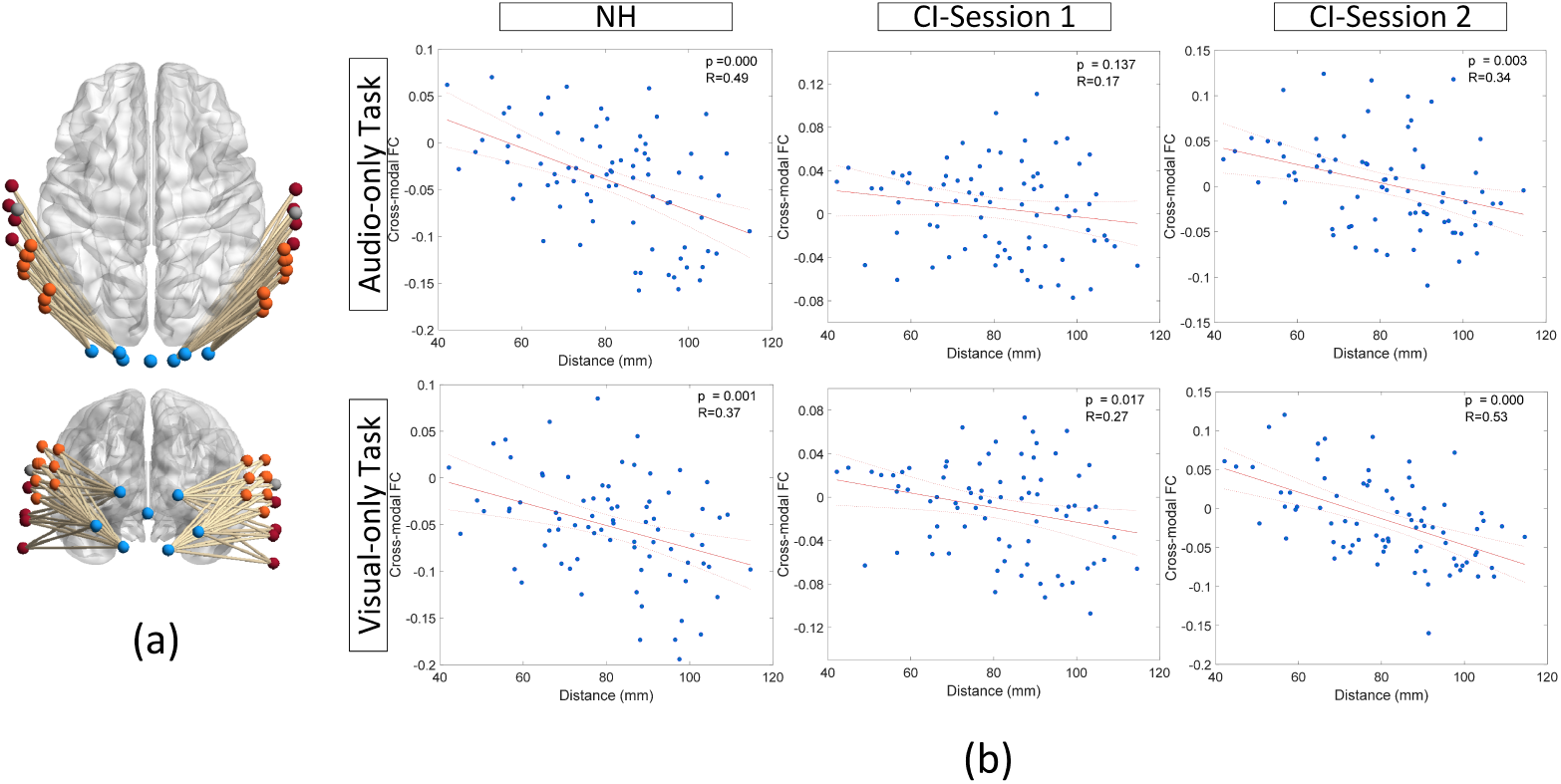
Correlation between task-induced CMFC and the distance between channel pairs within each hemisphere. Notably, for NH subjects, the correlation is negative and significant. Moreover, in the CI user group, the negative correlation strengthens after one year of device activation.

### 3.6. Task-induced CMFC of CI Users Compared to NH Subjects

We further investigated the task-induced CMFC of NH subjects and CI users. Fig.9 illustrates the distribution of average link weights across subjects. The results unveil a significantly higher mean value for task-induced CMFC in the CI group compared to NH participants at both time points—one month and one year after device activation.

**Figure 9:**
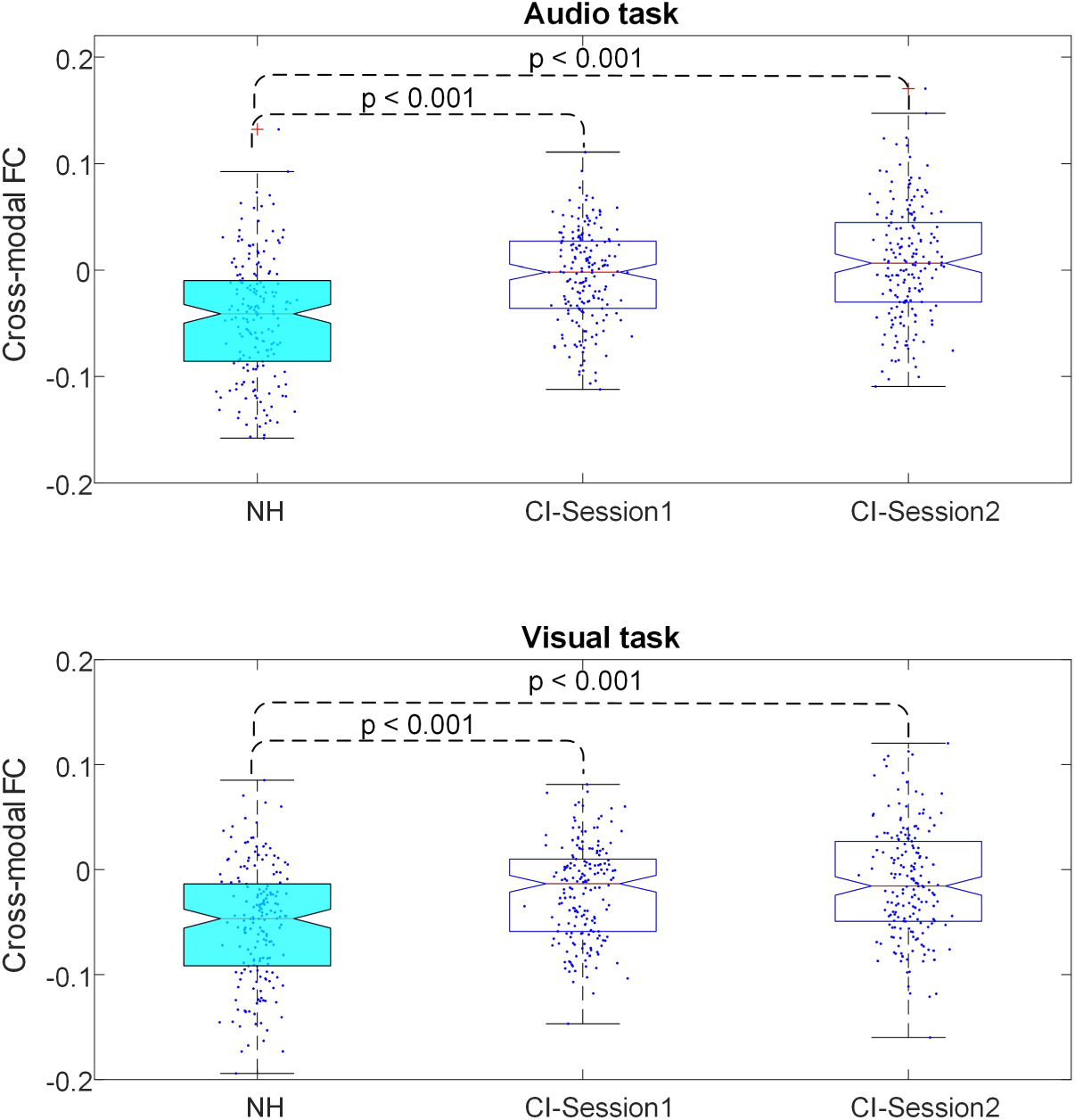
Comparison of the distribution of average task-induced CMFC across subjects for NH and CI groups. CI users exhibit significantly higher task-related CMFC values compared to the NH group (p-values represent the significance of the two-group comparison in each recording session). Dashed lines indicate two-group comparisons.

## 4. Discussion

Cross-modal plasticity underscores the brain’s remarkable capacity for reorganization in response to sensory input. However, several studies have highlighted the constraints and limitations of such reorganization, emphasizing the reliance on preexisting structural circuitry and top-down regulatory mechanisms [39, 40, 41]. This evidence aligns with the notion that the substrate of cross-modal plasticity primarily involves synaptic and functional adaptations, operating at the level of synaptic gain and inhibition, which subsequently lead to changes in functional connectivity without significant large-scale rewiring in the brain[5]. In line with this understanding, our study aimed to assess task-induced changes in CMFC between auditory and visual cortices in post-lingually deaf CI recipients, as a critical indicator of cross-modal plasticity following CI activation.

Functional brain imaging was conducted using fNIRS, a noninvasive optical imaging technique that measures changes in oxy- and deoxy-hemoglobin concentration within cortical blood flow. While fNIRS have lower spatial resolution compared to fMRI, its advantages including minimal noise, cost-effectiveness, and compatibility with implant devices make it particularly well-suited for hearing research [29, 42].

### 4.1. Task-induced CMFC Measurement Methodology

Drawing inspiration from Friston et al.’s Psychophysiological Interactions method [20], we devised an approach to quantify task-induced CMFC. There were two significant limitations inherent in the application of PPI, which we effectively addressed by utilizing closed-eyes resting-state recordings to establish baseline FC. Firstly, the generalized PPI method, typically employed for event-related experiments involving multiple tasks occurring randomly, lacks the ability to isolate the specific effects of individual tasks on FC, instead primarily contrasting the overall effects of multiple tasks [22, 43]. Secondly, we identified evidence suggesting that short control trials interspersed between task trials may be susceptible to contamination by residual task-related activity, with the brain’s FC potentially failing to return rapidly to baseline levels within these intervals [44]. Given these challenges, particularly relevant to our evaluation of the brain’s response to visual-speech tasks with participants maintaining open eyes during control trials, employing closed-eyes resting-state recordings for establishing baseline FC effectively resolved both issues, ensuring the accuracy and reliability of our baseline measurements while enabling the precise capture of task-induced CMFC changes.

### 4.2. Distinct Patterns of Cross-Modal Plasticity Revealed in Auditory Cortex Subregions

In our investigation of the correlation between task-related CMFC and speech performance outcomes among CI users, we initially focused on the broad areas within the auditory cortex encompassed by our montage setup. Our only precondition was the classification of cross-modal links into contra and ipsilateral categories relative to the implant side, reflecting the experimental conditions where participants received auditory stimuli through the cochlear implant on one side. Given the well-established lateralization patterns in structural auditory pathways [45], this distinction was deemed crucial. Our findings unveiled a notable negative correlation, with some instances reaching statistical significance (at *α* = 0.05), between average contra-lateral task-related CMFC observed in fNIRS recordings obtained in one month post-device activation and those taken after one year, and the speech understanding scores recorded after one year (Fig. 3).

Subsequently, we narrowed our focus to the restricted areas within the auditory cortex delineated by our previous research [29]. This analysis revealed significant differences between AG and STG concerning their correlation with the behavioral speech outcomes of the links originating from channels within these regions to VC. Specifically, our results indicated that the observed correlation, predominantly evident in the initial examination of broad areas, was largely driven by links originating from AG to VC (Fig. 4 and Fig 5). To confirm these disparities, we ranked all links and found notably higher significance scores for links originating from AG in terms of their correlation with speech scores (Fig. 6). Furthermore, our assessment of brain plasticity revealed a significant difference emerging after one year in task-related CMFC for links from AG to VC compared to those from STG to VC, with higher amplitude observed for AG links (Fig. 7).

These findings underscore a distinct divergence between AG and STG concerning cross-modal plasticity post-cochlear implantation, as assessed through task-induced CMFC. They emphasize the heightened significance of the AG area in facilitating cross-modal cooperation with VC, alongside the more pronounced plastic changes evidenced by significantly higher amplitude CMFC of links originating from this area to VC compared to links from STG to VC one year post switch-on. Thus, our results reaffirm studies indicating cross-modal activities after hearing loss predominantly relate to specific auditory regions, primarily associated with secondary auditory processing regions. These studies suggest that cross-modal supranormal enhancements of the visual modality are mostly restricted to enhanced visual motion detection and localization abilities associated with cortical auditory areas underlying localization and movement (in the AG area in our montage), but not all visual abilities like visual acuity and contrast sensitivity [5, 41, 46].

### 4.3. Link Length and Task-Induced CMFC: Insights into Brain Economy

One notable topological difference between AG and STG lies in their respective distances from VC. This prompted us to investigate whether link length, defined as the distance between pairs of channels, could influence task-induced CMFC. In essence, we sought to determine whether closer areas in AC and VC exhibit greater cooperation in task performance. Given the brain’s economic considerations, it’s reasonable to expect that shorter links would experience more pronounced task-related changes in FC. Neural elements and their connections are constrained by limited brain volume, making material and metabolic costs a significant concern. However, brain networks must also prioritize topological efficiency, robustness, and computational performance. Thus, the placement of neuronal components should ideally be optimized to balance these competing demands [47].

To explore this further, we examined the correlation between link length and task-related FC for links within each hemisphere in both CI users and NH subjects. As the physical connections between nodes in VC and AC located in different hemispheres are not straightforward and Euclidean distance may not be a proper estimation, we focused on links within each hemisphere. Our results (Fig. 8) demonstrated that, overall, in the NH group, task-related FC tends to decrease as the distance between nodes increases, showing a highly significant negative correlation. Shortly after implantation, CI users exhibited a weaker negative correlation, but after one year, a similar trend emerged. This suggests that the brain requires time to adapt to the new stimulation provided by the implant device, as indicated by user performance, where most changes occur within the first six months and stabilize after one year. These findings, in line with the brain’s economy principle, imply that plastic changes related to CMFC after implantation primarily adhere to principles of neural efficiency, showing higher task-related CMFC correlation for links between closer channels [47].

### 4.4. Enhanced Task-Induced CMFC in Cochlear Implant Users

Upon comparison of task-induced CMFC during both visual-speech and audio-only tasks between NH individuals and CI users, a significant distinction emerged: CI users displayed a notably higher mean task-induced CMFC compared to the NH group (Fig. 9). This finding indicates a stronger correlation in cross-modal brain activities during tasks among CI users than among NH subjects. Essentially, CI users, similar to individuals with hearing loss, exhibit an amplified reliance on cross-modal activities to comprehend both audio and visual speech. This dependence might be attributed to the cochlear implant’s delivery of signals with coarse spectral content in contrast to the natural acoustic hearing experienced by NH individuals, coupled with structural changes in the hearing system following hearing loss [6, 7], which requires more pronounced CI users’ reliance on cross-modal activities even after one year of using the device.

## 5. Conclusion and Future Directions

This study explored cross-modal plasticity among CI recipients with severe hearing loss in the contralateral ear, focusing on task-induced CMFC. To do so, we proposed a robust method inspired by PPI while addressing its limitations. We found a negative correlation between average contralateral task-induced CMFC and speech outcomes, particularly pronounced in links originating from AG, both one month and one-year post-device activation. Further distinctions between primary and secondary auditory areas, specifically AG and STG, revealed plastic cross-modal changes in one year post-switch-on, resulting in higher task-induced CMFC in AG areas compared to STG, aligning with principles of neural efficiency. Task-induced CMFC remained elevated in CI users compared to NH cohorts even after one year post-implantation, suggesting a sustained reliance on cross-modal activities. Future studies might assess more extended time intervals beyond one year using the proposed approach in this study.

## Data Availability

All data produced in the present study are available upon reasonable request to the authors

## Acknowledgement

This study was supported by a grant from the National Health and Medical Research Council of Australia to Colette M. McKay. Tommy Peng was supported by a Junior Fellowship from the Passe and Williams Foundation. Jamal Esmaelpoor was supported by the University of Melbourne Research Scholarship. The Bionics Institute acknowledges the support it receives from the Victorian Government through its Operational Infrastructure Support Program.

## Footnotes

1 They named these areas Herschel’s gyrus and Plenum temporale in their study.

